# Humoral and cellular responses to SARS-CoV-2 vaccination in patients with lymphoid malignancies

**DOI:** 10.1101/2021.12.08.21266760

**Authors:** Sean H. Lim, Nicola Campbell, Beth Stuart, Marina Johnson, Debora Joseph-Pietras, Adam Kelly, Danielle Jeffrey, Anna H. Turaj, Kate Rolfvondenbaumen, Celine Galloway, Thomas Wynn, Adam R. Coleman, Benjamin Ward, Karen Long, Andrew T. Bates, Diana Ayres, Robert Lown, Janlyn Falconer, Oliver Brake, James Batchelor, Victoria Willimott, Anna Bowzyk Al-Naeeb, Lisa Robinson, Ann O’Callaghan, Graham P. Collins, Tobias Menne, Saul N. Faust, Christopher P. Fox, Matthew Ahearne, Peter W.M. Johnson, Andrew J. Davies, David Goldblatt

## Abstract

SARS-CoV-2 vaccination protects against COVID-19. Antibodies and antigen-specific T-cell responses against the spike domain can be used to measure vaccine immune response. Individuals with lymphoma have defects in humoral and cellular immunity that may compromise vaccine response. In this prospective observational study of 457 participants with lymphoma, 52% of participants vaccinated on treatment had undetectable anti-spike IgG antibodies compared to 9% who were not on treatment. Marked impairment was observed in those receiving anti- CD20 antibody within 12 months where 60% had undetectable antibodies compared to 11% on chemotherapy, which persisted despite three vaccine doses. Overall, 63% had positive T-cell responses irrespective of treatment. Individuals with indolent B-cell lymphoma have impaired antibody and cellular responses that were independent of treatment. The significant reduction and heterogeneity in immune responses in these individuals emphasise the urgent need for immune response monitoring and alternative prophylactic strategies to protect against COVID- 19.

## INTRODUCTION

Individuals with cancer have increased morbidity and mortality from COVID-19 infection (Bange et al., 2021; Lee et al., 2020). This is most apparent in patients with haematological malignancies, who have a reported odds ratio of 1.57 to 3.3 in developing severe COVID-19, compared to patients with solid tumours (Bange et al., 2021; Lee et al., 2020). SARS-CoV-2 vaccination has been shown to be highly efficacious in preventing symptomatic COVID-19 disease in healthy individuals (Polack et al., 2020; Ramasamy et al., 2021). Recent data indicate that the presence of both binding and neutralising antibodies are highly predictive of protection against symptomatic disease, although a threshold correlating with protection is yet to be determined (Earle et al., 2021; Khoury et al., 2021). A number of observational studies have reported that whilst the majority of patients with solid malignancies develop anti-spike (S) IgG antibodies to vaccination, a significant proportion of patients with haematological malignancies, in particular those with lymphoid malignancies, do not (Addeo et al., 2021; Ehmsen et al., 2021; Greenberger et al., 2021b; Herishanu et al., 2021; Lim et al., 2021; Maneikis et al., 2021; Parry et al., 2021; Thakkar et al., 2021; Van Oekelen et al., 2021). In patients with lymphoid malignancies, a significant proportion of those who are vaccinated whilst receiving or recently completed B-cell depleting or targeted treatments such as anti-CD20 monoclonal antibodies and Bruton Tyrosine Kinase (BTK) inhibitors do not have detectable antibodies. In our initial interim analysis, we also reported that a proportion of individuals with chronic, indolent B-cell malignancies have impaired serologic responses regardless of relationship to the time of treatment (Lim et al., 2021).

Less well-explored is the role of vaccine-induced antigen-specific T cells in mediating protection, specifically a T-cell response to peptides expressing the SARS-CoV-2 S domain. Many of the SARS-CoV-2 vaccines induce demonstrable T-cell responses but due to the technical complexities of assessing a T-cell response, the majority of observational studies have not incorporated cellular responses after vaccination (Addeo et al., 2021; Greenberger et al., 2021b; Herishanu et al., 2021; Lim et al., 2021; Maneikis et al., 2021; Parry et al., 2021; Thakkar et al., 2021; Van Oekelen et al., 2021).

The other important factor in evaluating the immune response elicited by the vaccine is the functional quality of the antibodies produced. Anti-S antibodies can protect against SARS-CoV-2 infection by a number of different mechanisms which includes binding to the receptor-binding domain (RBD) within the S protein to sterically block its subsequent binding to host ACE2 receptor, and therefore viral entry into the cell (Yan et al., 2020). Although live virus neutralisation is the gold standard assay for assessing the presence of neutralising antibodies against SARS-CoV-2, this is a specialist assay with limited global capacity. Instead, pseudoneutralization assays which quantifies the ability of patient serum or plasma to inhibit the interaction between viral spike protein and soluble ACE2 receptor have been shown to be a useful surrogate for functional assessment (Goldblatt et al., 2021; Liu et al., 2021; Tan et al., 2020).

To address these points, we conducted a UK multicentre prospective observational study evaluating COVID-19 vaccine responses in individuals with lymphoma (PROSECO; NCT4858568) (Lim et al., 2021). Our first interim analysis reported antibody responses from 129 participants after two doses of either ChAdOx1 nCoV-19 (ChAdOx1) or BNT162b2 vaccines, given 10-12 weeks apart. We now provide an extended analysis involving 457 participants with lymphoma with detailed serological, cellular and pseudoneutralization responses.

## RESULTS

### Baseline characteristics

There were four hundred and fifty-seven participants with serological data. Twenty-nine participants had detectable anti-nucleocapsid (N) IgG antibodies, indicating prior SARS-CoV-2 infection and were excluded from the serological and cellular response analysis.

The baseline demographics, clinical characteristics and treatment details are described in Tables 1 and 2. Participants had Hodgkin lymphoma (HL, n=71), aggressive B cell non-Hodgkin lymphoma (B-NHL, n=149), indolent B-NHL (n=221) and peripheral T/NK cell lymphoma (PTCL, n=16). Participants with HL were younger than the other disease groups, with a median age of 40 years compared to 63 to 67 years, reflective of the presentation age of HL. In the aggressive B-NHL cohort, diffuse large B-cell lymphoma (DLBCL) was diagnosed in 83% (124/149) of participants. In the indolent B-NHL cohort, follicular lymphoma (FL) comprised 42.1% (93/221) of cases, followed by 23.1% (51/221) chronic lymphocytic leukaemia (CLL), 9.5% (21/221) mantle cell lymphoma, 9.5% (21/221) marginal zone lymphoma and 7.7% (17/221) lymphoplasmacytic lymphoma. The PTCL cohort was heterogeneous with angioimmunoblastic T-cell lymphoma comprising 43.8% (7/16) of the cohort. In each disease group, more patients received ChAdOx1 than BNT162b2 vaccines (overall: 60.4% (276/457) vs 39.4% (180/457), respectively).

**Table 1.**
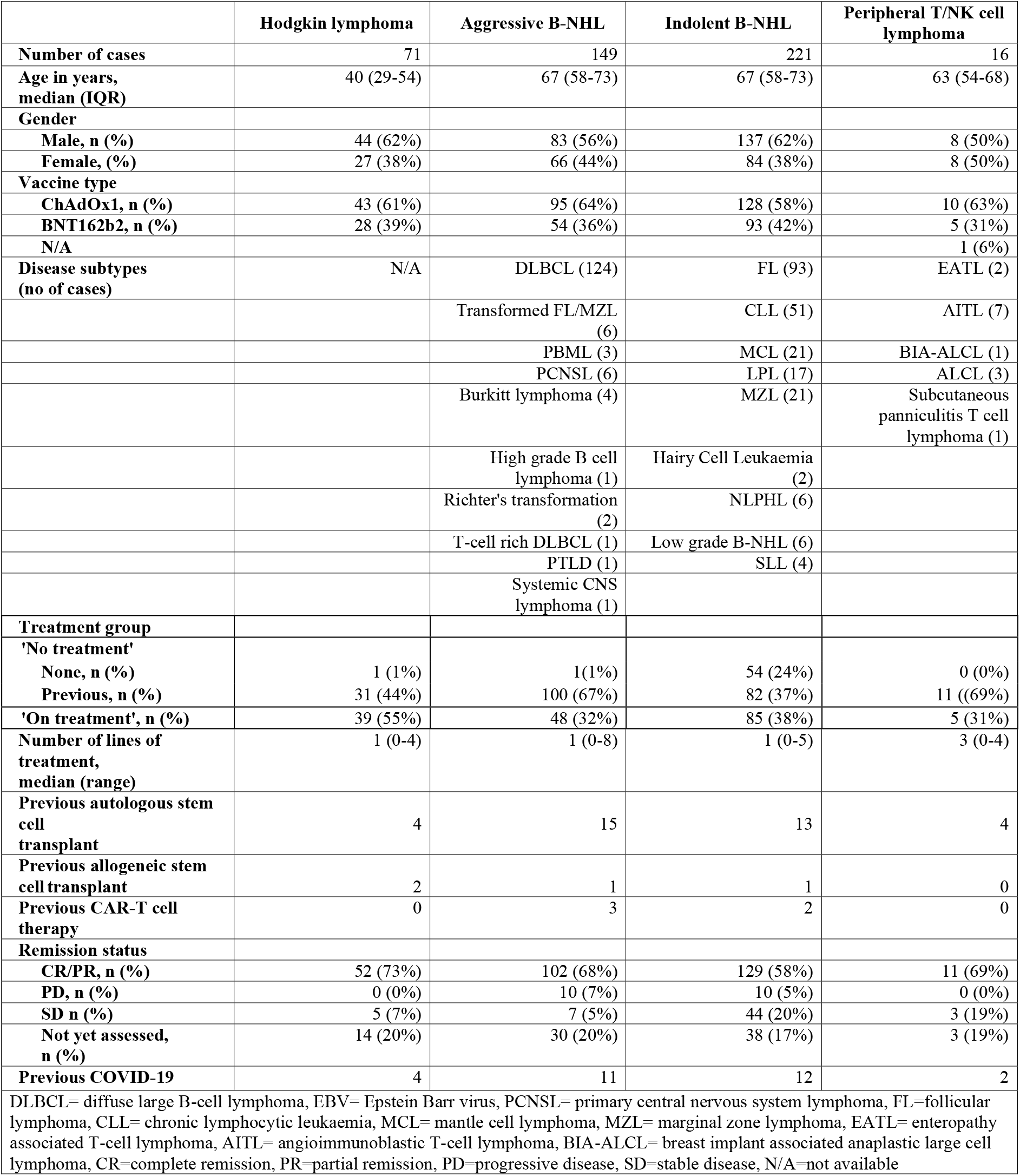
Baseline characteristics of participants

**Table 2.** Treatments received by participants within the ‘On Treatment’ group

| Last line of treatment | HL | Aggressive B-NHL | Indolent B-NHL | PTCL |
| --- | --- | --- | --- | --- |
| Chemotherapy | 27 | 0 | 0 | 1 |
| Anti-CD20 alone | 0 | 2 | 27 | 0 |
| Anti-CD20 & chemotherapy | 0 | 36 | 14 | 0 |
| Anti-CD20 & chemotherapy-free agent | 0 | 1 | 6 | 0 |
| Bendamustine | 1 | 2 | 0 | 0 |
| Bendamustine & anti-CD20 | 0 | 0 | 5 | 0 |
| BTK inhibitors | 0 | 0 | 17 | 0 |
| Venetoclax | 0 | 0 | 7 | 0 |
| BTK inhibitor & venetoclax | 0 | 0 | 2 | 0 |
| CAR-T cell therapy | 0 | 0 | 0 | 0 |
| Autologous stem cell transplant | 0 | 4 | 2 | 0 |
| Allogeneic stem cell transplant | 0 | 0 | 0 | 0 |
| Others | BV (2),<br>BV +<br>Bendamustine (1) | CPI (1)<br>Bispecific Ab (1) | Bispecific Ab +<br>lenalidomide (1) | PI3K inhibitor (1)<br>BV (1)<br>ciclosporin (1) |
| BV=brentuximab vedotin, CP=checkpoint inhibitor, PI3K=phosphatidylinositol 3-kinase |  |  |  |  |

Overall, 38.7% (177/457) participants in the ‘on treatment’ group, which was defined as those who received their first vaccine dose within 24 weeks of completing systemic anti-lymphoma treatment, or whose treatment had commenced within 4 weeks of receiving the first vaccine dose. Forty-nine per cent (224/457) of participants had completed their treatment more than 24 weeks before their first vaccine dose and were allocated to the ‘no treatment’ group. A further 12.3% (56/457) participants had not received systemic treatment for their disease prior to the first vaccine dose; comprising patients with indolent B-NHL (96.4%, 54/56) who were also allocated to the ‘no treatment’ group.

At the last recorded visit, 64.3% (294/457) were in complete or partial remission, 12.9% (59/457) had stable disease and 4.4% (20/457) had progressive disease. A further 18.2% (83/457) participants’ remission status had not been evaluated as they were continuing to receive treatment.

Anti-S, anti-RBD and anti-nucleocapsid (N) IgG concentrations were measured in 56, 67 and 430 participants prior to vaccination (TP00), 4 weeks post first dose (TP01), and 2 to 4 weeks post second dose of vaccine (TP02), respectively (Figure 1). Thirty-two participants had cellular responses investigated by paired T-cell IFNγ ELISpot testing on peripheral blood mononuclear cells (PBMCs) collected at TP00 and TP02. A further 159 participants had ELISpot testing on TP02 only. Peripheral blood T, B and NK cell counts were also simultaneously performed alongside ELISpot testing. The ACE2 receptor blocking assay was performed on 282 samples at TP02. Serum was available from 136 healthy donor volunteers for serological analysis, and a further four and six donors for ELISpot and T, B and NK analysis, respectively.

**Figure 1.**
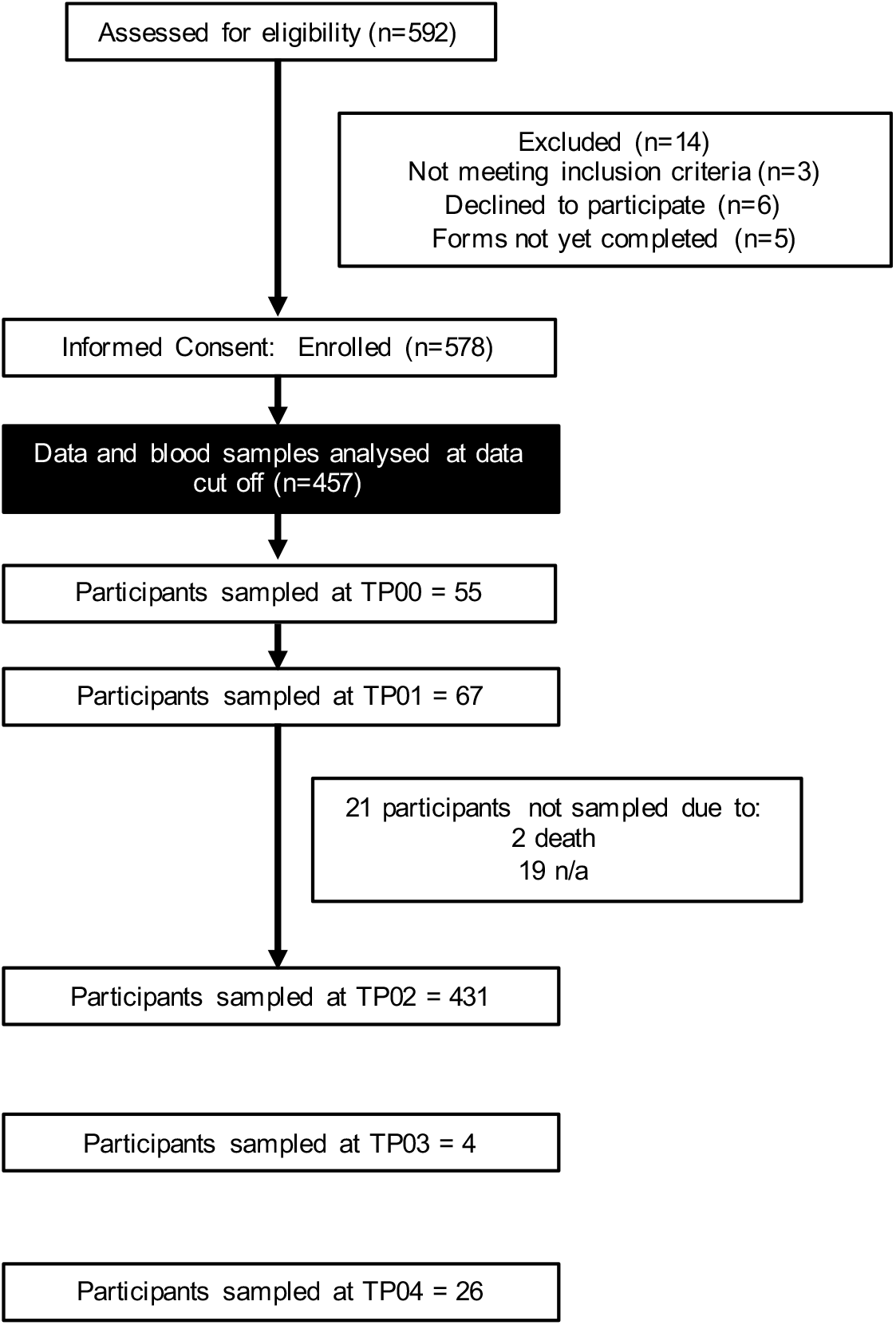
CONSORT diagram of study cohort. Five hundred ninety-two patients were assessed for eligibility and subsequently 578 recruited into the study. At the time of data cut-off, blood samples were processed from 457 participants. Numbers of participants sampled at the following time points are shown: pre-vaccination (TP00), 4 weeks post first dose (TP01), 2 to 4 weeks post second dose (TP02), 24 weeks post second dose (TP03) and 4 to 8 weeks post third dose (TP04).

### Patients vaccinated on treatment have impaired antibody responses but preserved cellular responses

The lymphoma participants were analysed separately, depending on whether they received BNT162b2 or ChAdOx1 vaccines (Figure 2A and B). The proportion of participants who had undetectable antibodies after two doses of each vaccine was similar in the ‘no treatment’ group (ChAdOx1 (9.1%, 14/154) vs BNT162b2 (8.2%, 8/98)). Whilst participants who had received two doses of BNT162b2 achieved a 2.4-fold higher antibody level than ChAdOx1 recipients, this was not statistically significant (geometric mean concentration (GMC) 270.8 (95% CI 156.9, 467.6) vs 111.5 (78.35, 158.7) BAU/mL, Kruskal-Wallis test). In healthy donors, BNT162b2 induced higher anti-S IgG antibody levels than ChAdOx1, as observed in our earlier analysis (Lim et al., 2021). The GMC of healthy donors vaccinated with two doses of BNT162b2 was 11-fold higher than ChAdOx1 (2667 vs 196 BAU/mL, p=0.0043, Kruskal-Wallis test).

**Figure 2.**
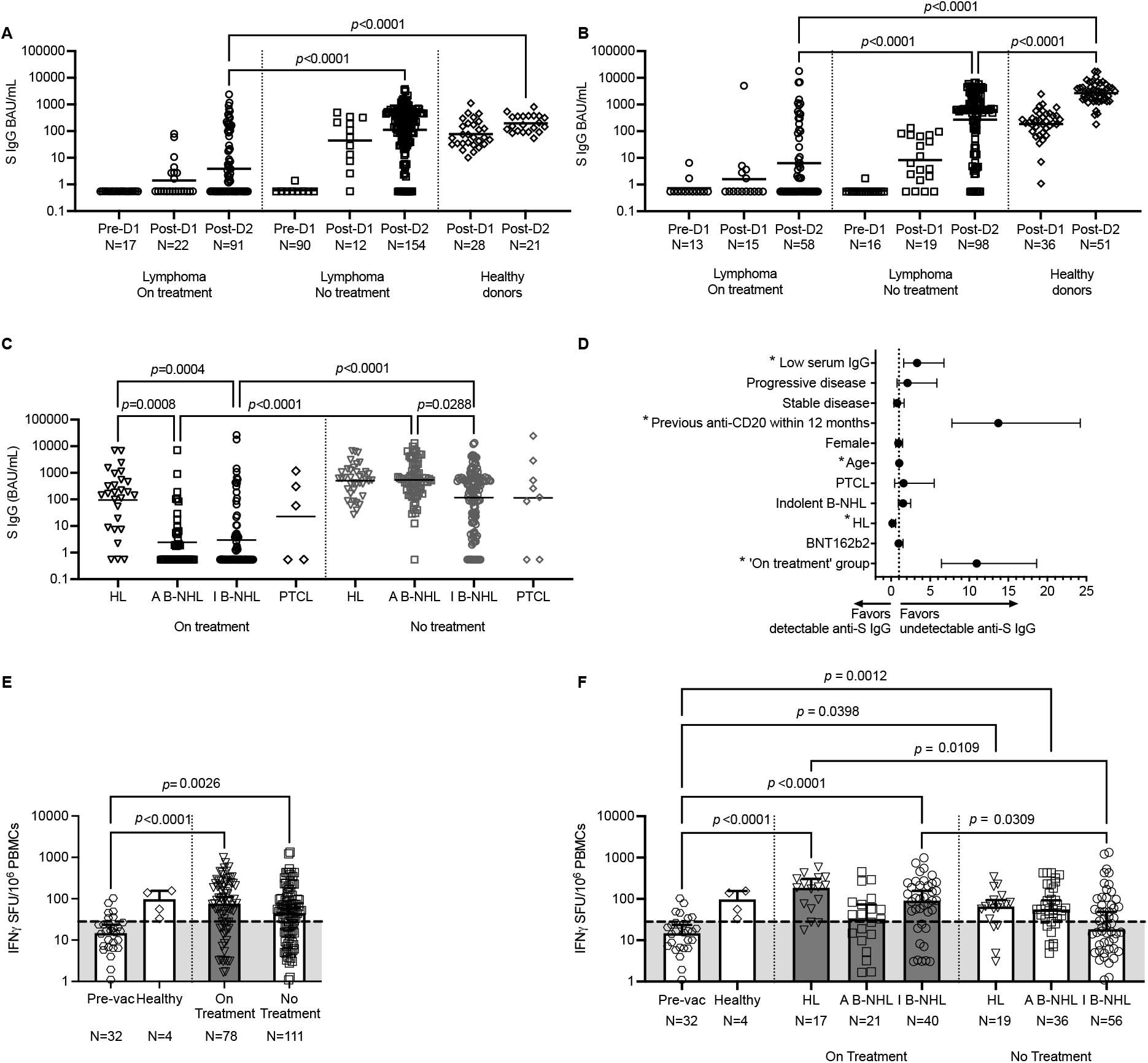
Antibody responses but not cellular responses to SARS-CoV-2 vaccination are impaired by systemic therapy. (A) Lymphoma participants vaccinated with ChAdOx1 vaccine whilst on systemic treatment or within 24 weeks of completion had significantly less detectable anti-S IgG antibodies and reduced antibody levels compared to previous or untreated lymphoma participants and healthy controls. GMC shown, Kruskal-Wallis test. (B) As in (A) but with BNT162b2 where the same phenomenon is observed. (C) Reduced anti-S IgG antibody levels in HL, aggressive B-NHL (A B-NHL) and indolent B-NHL (I B-NHL) participants in the ‘on treatment’ group, and in I B-NHL participants in the ‘no treatment’ group despite two vaccine doses. GMC shown, Kruskal-Wallis test. See Figure S2. (D) Univariable logistic regression showing that low serum IgG, anti-CD20 exposure 12 months preceding first vaccine dose, ‘on treatment’ group and older age were associated with an increased OR of having undetectable anti-S IgG. HL favoured detectable anti-S IgG. *marks statistical significance. See Table S1. (E) Similar IFNγ cellular responses in lymphoma participants between ‘on’ and ‘no treatment’ groups. Median and 95% CI shown, Kruskal-Wallis test. (F) Reduced IFNγ cellular responses were preserved in lymphoma participants except for those with I B-NHL in the ‘no treatment’ group.

Participants in the ‘on treatment’ group had impaired antibody responses compared to ‘no treatment’, regardless of vaccine type. Overall, 52.3% (78/149) of participants ‘on treatment’ had undetectable antibodies after two doses of vaccine compared to 8.7% (22/252) in the ‘no treatment’ group. Given that there was not a statistically significant difference in antibody levels and the proportion of undetectable antibodies was similar between the two vaccines in participants with lymphoma, ChAdOx1 and BNT162b2 groups were merged for subsequent analysis.

Within the ‘on treatment’ group, fewer participants with HL had undetectable antibodies compared to aggressive and indolent B-NHL (11.1%, (3/27) vs 56.8% (25/44) vs 62.7% (47/78), respectively) (Figure 2C). The sample size of the PTCL group was too small to comment on treatment effect. Overall 30.7% (4/13) of participants with PTCL had no detectable antibodies. Consistent with Figure 2A, more participants in the ‘no treatment’ group had detectable antibodies. Specifically, 100% (38/38) of HL and 97.7% (86/88) of aggressive B-NHL cases had detectable antibodies and 90.6% (106/117) of participants with indolent B-NHL. The anti-S level was similar for HL and aggressive B-NHL but reduced for indolent B-NHL (GMC 502.9 vs 539.1 vs 116.5 BAU/mL, p=0.0288, Kruskal-Wallis test). In those with indolent B-NHL, there was no significant difference in the antibody level between participants who had never received systemic therapy for their disease versus those who were previously treated, suggesting that participants with indolent B-NHL may have additional treatment unrelated disease-intrinsic immune dysfunction that impairs antibody levels (Figure S2).

A univariable analysis showed that participants in the ‘on treatment’ group (odds ratio (OR) 10.96), anti-CD20 administration in the last 12 months (OR 13.72), low serum IgG level (OR 3.32) and increasing age (OR 1.04) was were associated with an increased risk of having no detectable anti-S IgG (Figure S2D, Table S1). On multivariable analysis, ‘on treatment’ group (OR 7.22) and anti-CD20 administration in the last 12 months (OR 5.60) continued to be significant factors (Table S2). Serum IgG was not included for multivariable analysis as a result was available for only 210 participants, which would have substantially reduced the statistical power of the analysis.

The antigen-specific T cell response in the lymphoma group, as analysed by IFNγ ELISpot assay using peripheral blood mononuclear cells (PBMCs), showed no significant difference between ‘on’ and ‘no treatment’ groups (median 76.2 IFNγ spot forming units (SFU)/10^6^ PBMCs (95% CI 50.8, 110.0) vs 45.10 (27.97, 57.13), p=0.16, Kruskal-Wallis) (Figure 2E). However, there was a variation in cellular response within the disease groups (Figure 2F). In HL, the proportion of positive cellular responses was equivalent between ‘on’ and ‘no’ treatment groups (75% (12/16) vs 73.9% (17/23)). In contrast, 52% (11/21) of participants with aggressive B-NHL who were ‘on treatment’, had a positive cellular response compared to 76.5% (26/34) of participants in the ‘no treatment’ group although there was no statistical difference in the actual number of IFNγ SFU/10^6^ PBMCs between the two groups. In indolent B-NHL, those in the ‘no treatment’ group had an inferior response compared to those on treatment (positive responses: 44.6% (25/56) vs 72.5% (29/40)) and this was reflected by a significant difference in the number IFNγ SFU/10^6^ PBMCs (no vs on treatment: median 18.47 vs 91.09, p=0.0309, Kruskal-Wallis).

### Anti-CD20 therapy-related serologic impairment can be predicted by peripheral blood B cell counts

It is recognised that anti-CD20 monoclonal antibody therapy profoundly suppresses the antibody response to SARS-CoV-2 vaccination. To ensure that the impairment in antibody response is not confounded by disease-related immune dysfunction, only participants with aggressive B-NHL were analysed for the impact of anti-CD20 (n=94) (Figure 3A). Fifty per cent (9/18) and 61.5% (8/13) of participants who received their first vaccine dose whilst on anti-CD20, or within 6 months of completion, respectively, had undetectable antibodies. Responses improved as the duration between completion of anti-CD20 prior to vaccination increased. One of four participants had undetectable antibodies when anti-CD20 had completed 7-12 months pre-vaccination. All participants who were vaccinated more than 12 months after anti-CD20 completion had detectable anti-S IgG (GMC 492.5 BAU/mL (95% 98.96, 2609)).

**Figure 3.**
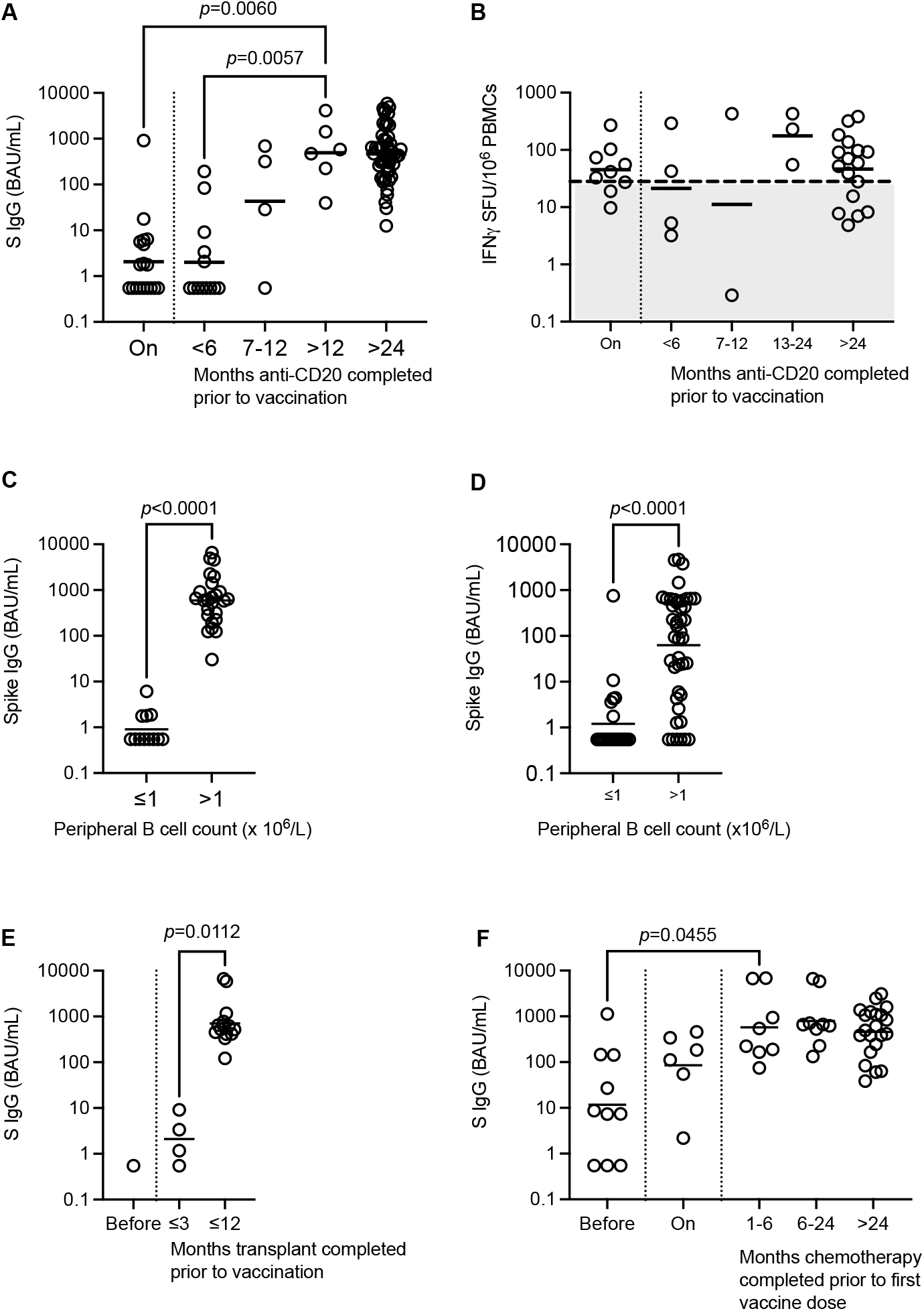
The effect of specific therapies on antibody and cellular responses. (A) Anti-S IgG levels in participants vaccinated whilst receiving anti-CD20 or <6 months of completion. GMC shown, Kruskal-Wallis test. (B) Preservation of cellular responses in anti-CD20-treated lymphoma participants, regardless of treatment timing. All points below the dashed line represents a negative test. Medians shown, Kruskal-Wallis test. (C) A reduced peripheral blood B-cell count is associated with impaired anti-S IgG level in aggressive B-NHL. GMC shown, Mann-Whitney test. (D) As in (C) but for indolent B-NHL. (E) Anti-S IgG levels in participants vaccinated before or within 3 months of autologous stem cell transplant have reduced anti-S. GMC shown, Kruskal-Wallis test. See Figure S3. (F) Participants with HL have reduced anti-S when vaccinated 3 months before commencing chemotherapy. See Figure S3.

With regards to T-cell response, a positive response was observed in 62.5% (10/16) and 71.4% (15/21) participants who had completed anti-CD20 less than 12 and 12 months or more prior to vaccination, suggesting that cellular responses are often preserved with anti-CD20 treatment (Figure 3B).

The antibody impairment mediated by anti-CD20 therapy is very likely to be related to B-cell depletion. Therefore, we compared the antibody responses of participants with a peripheral blood B cell count of ≤1 or >1 × 10^6^/L, within aggressive B-NHL (Figure 3C). There was a marked and significant difference in antibody level between those with B cell count ≤1 vs >1 (0.90 vs 590.1 BAU/mL, p<0.0001, Mann Whitney). For indolent B-NHL, a B-cell count exceeding 1 × 10^6^ cells/L was also associated with a higher antibody level but 13.6% (6/44) of participants in this group still had undetectable antibodies, suggesting that other factors beyond B-cell depletion contribute to impairment of vaccine antibody response (Figure 3D).

### Antibody responses are impaired if participants are vaccinated before or early post autologous stem cell transplant

The impact of stem cell transplantation on vaccine immune responses was examined by analysis of individuals whose last line of treatment was an autologous stem cell transplant (Figure 3E). Here, 5/19 participants who had received two doses of vaccine either 3 weeks before transplantation, or within 5 months after transplantation, had a reduced antibody response compared to those vaccinated 12 months or more after transplantation (GMC 1.61 BAU/mL (95% CI 0.35, 7.31) vs 707 BAU/mL (95% CI 384.4, 1300)). T-cell ELISpot was undertaken in 8 participants who were transplanted more than 12 months ago, and 3/8 participants had a negative response (data not shown). Peripheral T, B and NK counts were within the normal ranges for both (data not shown). There were four participants who had received an allogeneic stem cell transplant more than 2 years prior to vaccination (Figure S3A). All had detectable antibodies and cellular response was observed in the one participant tested.

### Individuals with Hodgkin Lymphoma develop robust antibody and detectable cellular responses

In order to assess the effect of chemotherapy on vaccine immune responses without the contribution of anti-CD20, we focused on a cohort of 54 participants with HL who had TP02 antibody levels (Figure 3F). Three out of ten participants who were vaccinated within 12 weeks of starting chemotherapy had undetectable antibody responses compared to 1/6 participants who were vaccinated on chemotherapy. A lower antibody level was observed in the former but this was not statistically significant (GMC 11.7 BAU/mL (95% CI 1.78, 77.7) vs 85.18 BAU/mL (95% CI 11.0, 659.0)). However, those who were vaccinated prior to chemotherapy commencement had significantly lower antibody levels compared to those who were vaccinated 1 month or more after treatment completion (GMC 11.7 BAU/mL (95% CI 1.78, 77.7) vs 547.5 BAU/mL (95% CI 351.5-852.8); p<0.01, Kruskal-Wallis test).

Cellular responses were also examined in 31 participants with HL. No correlation was observed between antibody and cellular responses and the proportion with positive cellular response was similar irrespective of the time of treatment (vaccination prior to and on chemotherapy 79% (11/14) vs 76% (13/17) vaccinated more than a month after chemotherapy (Figure S3B).

Altogether, our data suggests that cellular responses are preserved in the majority of participants on chemotherapy in HL. Antibody responses are impaired when individuals are vaccinated shortly before (3 months) or during chemotherapy compared to those who are vaccinated after treatment completion.

### Cellular responses to SARS-CoV-2 vaccination does not correlate with antibody responses in lymphoma

Predictors of cellular response to SARS-CoV-2 vaccination are poorly described compared to serological responses. Consistent with the preceding data shown here within the specific treatment groups (Figure 2E and S3E), no correlation was observed between cellular responses and anti-S IgG levels across all 191 individuals analysed (Figure 4A). ChAdOx1 vaccination resulted in higher T-cell responses than BNT162b2, producing a 2.1-fold higher response (median 69.7 IFNγ SFU/10^6^ PBMCs (95% CI 55.45, 97.50) vs 33.25 (16.68, 56.28) respectively, p=0.0005, Mann-Whitney) (Figure 4B). In a multivariable analysis, vaccine type continued to be a statistically significant factor, with ChAdOx1 recipients having an increased odds ratio of a positive cellular response (OR 2.01, 95% CI 1.06, 3.79, p=0.031) (Figure S4A, Table S3). No correlation was observed between cellular response and age, gender and remission status or across disease groups.

**Figure 4.**
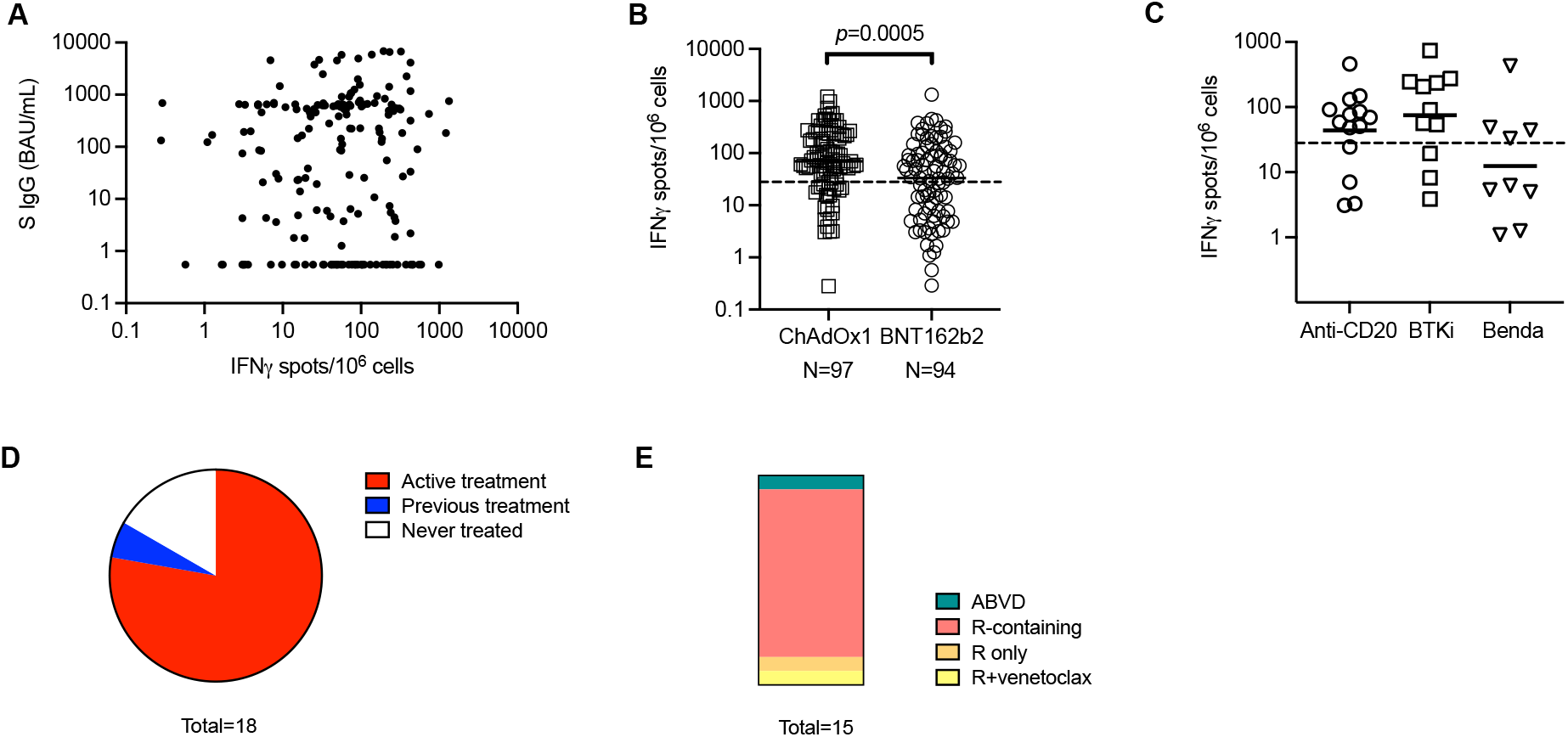
Correlates of antigen-specific T-cell responses. (A) Lack of correlation between antigen-specific IFNγ T-cell responses and anti-S levels. See Figure S4. (B) ChAdOx1 vaccination induces higher IFNγ T-cell responses than BNT162b2. All points below the dashed line represents a negative test. Medians shown, Mann-Whitney test. (C) Bendamustine treatment shows a trend towards reduction of IFNγ T-cell responses. All points below the dashed line represents a negative test. Medians shown, Kruskal-Wallis test. See Figure S4. (D) Most participants with negative antibody and cellular responses are from the ‘on treatment’ group. (E) Majority of the participants from the double negative, ‘on treatment’ group, receive combination therapy containing anti-CD20 rituximab (R).

Purine analogue-like chemotherapies such as bendamustine have previously been shown to reduce peripheral blood CD4^+^ T cells beyond 3 years after completion of treatment, thereby potentially impairing cellular responses (Marcus et al., 2017; Martinez-Calle et al., 2019). We compared participants with indolent B-NHL who were receiving, or had received bendamustine-containing regimens as the last line of regimen to those treated with anti- CD20 or BTK inhibitors(Figure 4C). Forty-four per cent (4/9) of bendamustine-treated participants had a positive response compared to 71.4% (10/14) and 81.8% (9/11) of anti-CD20 and BTK inhibitor-treated participants. However, no significant difference was observed in the absolute CD4^+^ T cell counts subset counts between the three groups, or between positive and negative responders within the bendamustine cohort (Figure S4B, C). Altogether these data suggest that unlike serological responses, systemic therapy is a poor predictor of cellular response in lymphoid malignancies. T-cell depleting therapies like purine analogues may impair cellular response but a larger cohort is required to confirm this observation.

### Factors determining a double negative immune response to vaccination

Theoretically, participants who have neither detectable antibodies nor cellular responses may be at greatest risk of a severe outcome to COVID-19 infection. In our cohort, 191 participants had paired serological and cellular response data 2-4 weeks post second vaccination. Nine percent (18/191) of participants had undetectable anti-S IgG antibodies and a negative cellular response (Figure 4D). Seventy-eight percent (14/18) participants were from the ‘on treatment’ group. Amongst these 14 participants, 8 had indolent B-NHL, 5 had aggressive B-NHL and 1 had HL. Apart from the participant with HL, all were receiving anti-CD20 containing chemoimmunotherapy (Figure 4E). In the remaining 4/18 participants from the ‘no treatment’ group, 3 were participants with indolent B-NHL who had never received systemic treatment for their disease and a further participant was previously treated with a purine analogue containing regimen (FCR: fludarabine, cyclophosphamide and rituximab) 7 years ago.

To provide perspective for these data, 10.3% (8/78) of participants with indolent B-NHL, 11% (5/44) with aggressive B-NHL and 3.7% (1/27) with HL from the ‘on treatment’ group have no cellular or antibody response to two doses of vaccine. Seven percent (4/56) of participants with indolent B-NHL in the ‘no treatment’ group also had undetectable cellular or antibody response. All of the 36 and 20 participants with aggressive B-NHL and HL in the ‘no treatment’ group had either a detectable antibody or cellular response.

### Anti-spike IgG antibody levels correlate with virus pseudoneutralization

We also evaluated the correlation between anti-S IgG level and functional ability of the antibody to block ACE2 to wild type spike protein using the ACE2 receptor inhibition assay (Figure 5A, B). A good agreement was observed between anti-S IgG level and ACE2 receptor inhibition in 282 lymphoma cases tested (r=0.93, p<0.0001, Spearman correlation test). However, at lower antibody concentrations (<52.6 BAU/mL), the correlation between anti-S IgG level and ACE2 receptor inhibition is reduced (r=0.29, p<0.0004). Accordingly, when the ACE2 receptor inhibition levels are arbitrarily cut off at 1 or less, 10 or less and more than 10, the anti-S IgG GMC level increases from 8.9 (95% CI 5.8, 12.8) to 242.6 (95% CI 196.3, 299.7) to 2546 (95% CI 2042, 3174). Altogether, this suggests that similar to healthy individuals, there is a good correlation between antibody level and neutralizing activity in patients with lymphoid malignancies (Goldblatt et al., 2021).

**Figure 5.**
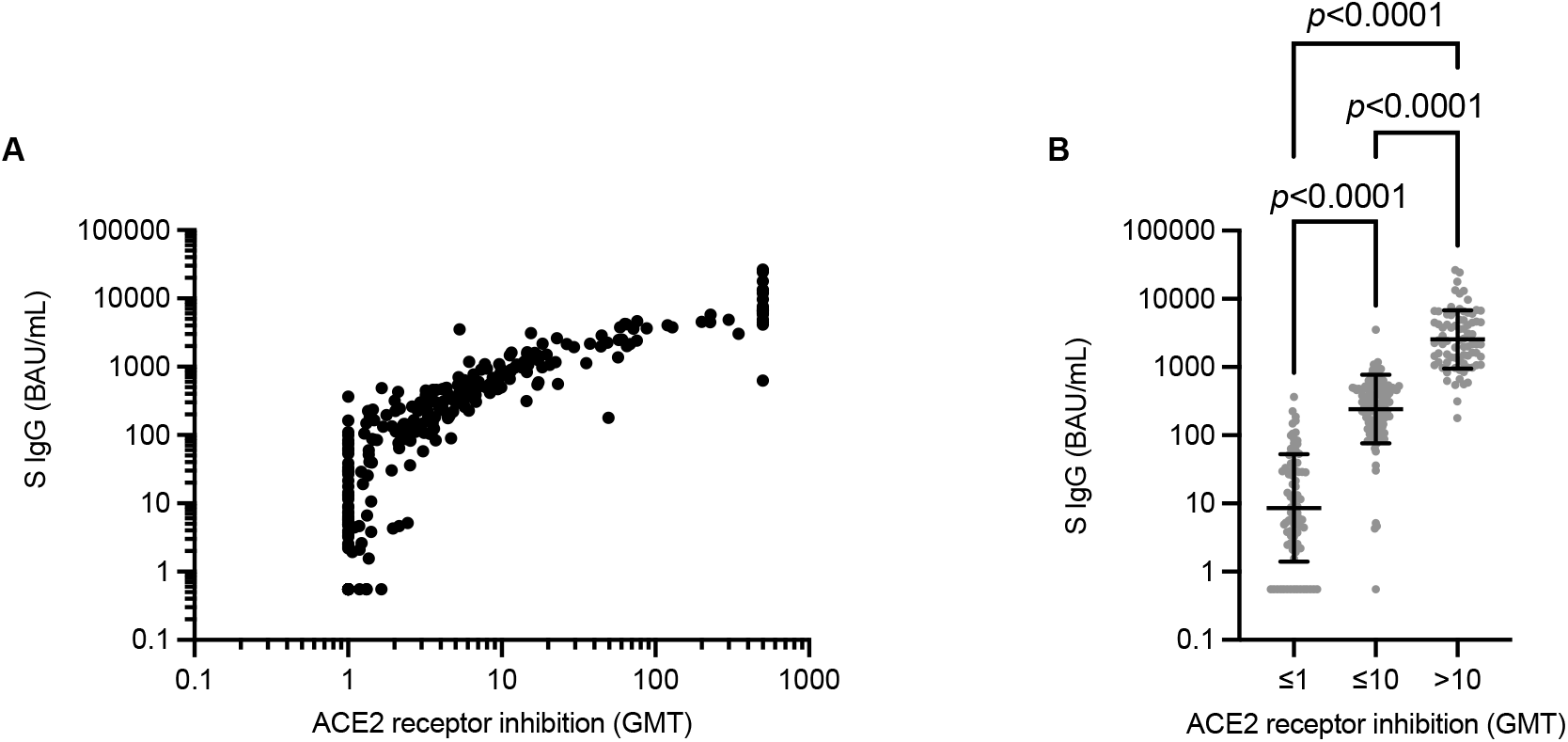
Anti-S level correlates with ACE2 receptor inhibition against WT SARS-CoV-2. (A) Correlation between anti-S IgG and ACE2 receptor inhibition. r=0.94, p<0.0001, Spearman test. (B) Data from (A) but divided according to ACE2 receptor inhibition levels. Medians shown, Kruskal-Wallis test.

### Third-dose vaccination within 12 months of anti-CD20 administration does not boost antibody responses

Twenty-six of the participants in the study had received a third vaccine dose 12 to 27 weeks after their second dose (Figure 6A). Four participants did not have a sample taken prior to the third vaccine dose. Eleven of the participants received their third vaccine dose within 52 weeks of anti-CD20 administration and 64% (7/11) had no detectable antibodies. In the remaining 4/11 patients, 3 had an antibody level below 10 BAU/mL and 1 participant, 78 BAU/ml. Two participants received the third vaccine dose whilst undergoing combination chemotherapy for HL. Both had detectable antibodies pre-vaccination and a modest increase was observed post vaccination (20.1 to 23.7; 36.5 to 60.9 BAU/ml). Amongst participants who had received any systemic treatment in the last 6 months, 6, 4 and 2 participants had indolent B-NHL, HL and aggressive B-NHL, respectively. Two of six FL participants had undetectable antibodies prior to third vaccine dose which was detectable but still low post vaccination (8.2 and 24.7 BAU/mL). The antibody level of three of four FL participants with detectable antibodies prior to the third vaccine dose rose by 7- to 24-fold. All 6/6 HL and aggressive B-NHL participants off treatment had detectable antibodies prior to the third vaccine dose, and there was a median 10.4-fold increase in antibody level after receipt of the third dose (range 2.0 to 36.0). Consistent with the anti-S IgG level, the lowest ACE2 receptor inhibition activity is observed in participants who received their third vaccine dose within 52 weeks of anti-CD20 administration (Figure 6B).

**Figure 6.**
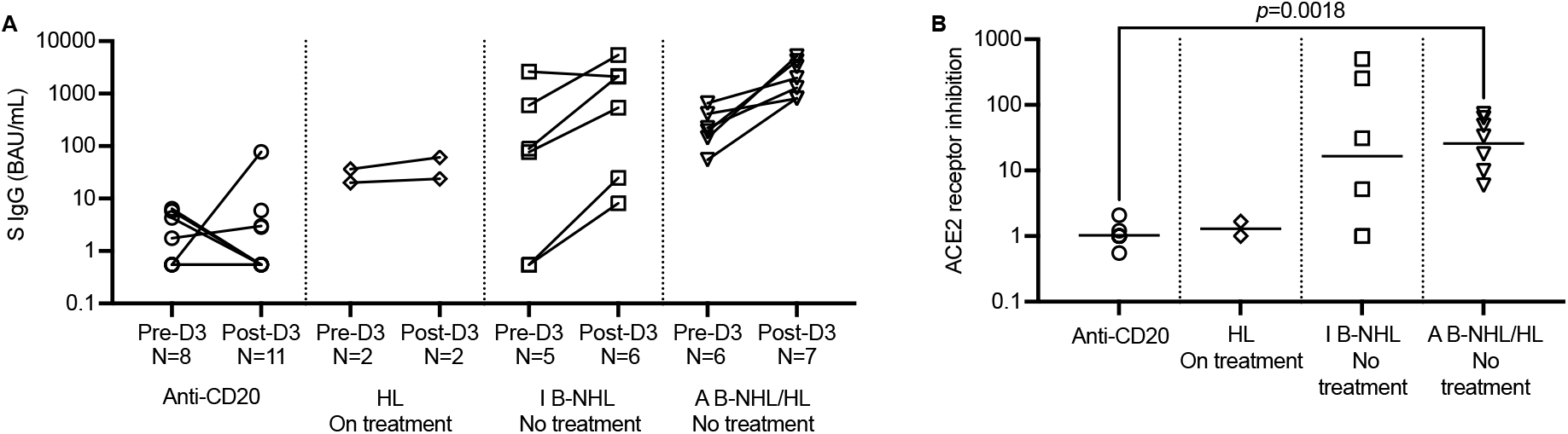
Antibody and pseudoneutralization responses to third vaccine dose. (A) Antibody responses to a third vaccine dose is impaired by ongoing or anti-CD20 therapy within 6 months in B-NHL and ongoing chemotherapy in HL. (B) TP03 matching pseudoneutralization results to (A).

## DISCUSSION

Our prospective cohort study evaluating antibody and cellular responses in 457 participants with lymphoid malignancies show that the timing of SARS-CoV-2 vaccination in relation to treatment is the strongest predictor of antibody response. No antibodies were detected after two vaccine doses, in 52.3% (78/149) of participants who were vaccinated within 24 weeks of completing their treatment compared to 8.7% (252) who had not received any systemic treatment within 24 weeks, after. Other than timing of treatment in relation to vaccination, the best predictors of antibody response were the peripheral blood B-cell count and serum immunoglobulin levels, consistent with data reported in autoimmune rheumatic disease.(Moor et al., 2021)

Anti-CD20 therapy markedly impaired antibody responses for at least 6 months post treatment. However, the preservation of antigen-specific T-cell responses may justify the continued vaccination of these patients, particularly if they are on a prolonged course of rituximab therapy. Other B cell-directed therapies such as CD19 CAR-T cell therapy, BTK inhibitors, ibrutinib and acalabrutinib, and Bcl2 inhibitor venetoclax also impaired antibody responses but not T-cell responses in our cohort but the numbers of patients were too small to reach any definitive conclusion (data not shown, (Lim et al., 2021)) However, other studies have also reported antibody impairment with these therapies (Herishanu et al., 2021; Parry et al., 2021).

Chemotherapy administered without anti-CD20, also impairs antibody responses, but to a far lesser extent. This is reflected by only 11% (3/27) of chemotherapy recipients having undetectable antibodies compared to 60% (58/96) in those receiving anti-CD20. However, the caveat being that this population largely comprised of patients with HL who tend to be younger than those with B-NHL. The majority of participants with other tumours types treated with chemotherapy also tend to mount antibody responses to the vaccine (Greenberger et al., 2021b; Waldhorn et al., 2021).

Data on cellular responses post SARS-CoV-2 vaccination are limited and where reported, most cohorts tend to be relatively small or heterogeneous in disease type, with the exception of Ehmsen and colleagues who analysed 323 participants, mostly comprising chronic lymphocytic leukaemia and multiple myeloma (Ehmsen et al., 2021; Fendler et al., 2021; McKenzie et al., 2021). There is an increasing body of data demonstrating that T cells can protect individuals with impaired antibody responses to COVID-19, or post-vaccination, against severe infection(Apostolidis et al., 2021; Bange et al., 2021). Consistent with other studies, we detected T-cell responses in participants on anti-CD20(Fendler et al., 2021). Due to the size of our dataset, we were also able to ascertain that systemic therapy had minimal effect on T-cell responses. One significant finding is the observation of anti-S T-cell responses in patients with HL regardless of treatment. It has long been suggested that these patients have a defect in their cellular immunity but our data shows no evidence of excess functional T-cell impairment when compared to B-NHL. (Foukaneli et al., 2020; Poppema, 1996)

Reduced T-cell responses were observed in participants with indolent B-NHL who had not recently had systemic treatment, compared to aggressive B-NHL and HL. T-cell dysfunction has been previously reported in FL and CLL so this is not entirely unexpected (Arruga et al., 2020; Ramsay et al., 2009). Further, Wherry et al also recently demonstrated elevated CD8^+^ T-cell responses in patients with multiple sclerosis treated with anti-CD20 (Apostolidis et al., 2021). One of the mechanisms hypothesised was that anti-CD20 may deplete regulatory B cells leading to loss of suppression of T cells. It could be that this phenomenon is apparent in indolent B-NHL and not aggressive B-NHL due to “lower baseline” T cell function. We also observed reduced cellular responses in participants treated with bendamustine, raising the possibility that participants treated with bendamustine and rituximab, a combination frequently employed in indolent B-NHL, may result in impairment of both antibody and cellular responses, and consequently, no protection against COVID-19. Unfortunately, the cohort is too small to draw any firm conclusions.

Our study has several limitations, first is that we did not assess the efficacy of the antibodies against SARS-CoV- 2 variants of concern. However, these data can be extrapolated from our accompanying publication, which reports the efficacy of these antibodies, measured in the same laboratory, against the variants (Goldblatt et al., 2021). The good agreement between antibody level and ACE2 receptor blocking against the wild type spike protein, suggests that these patients have functionally similar antibodies to healthy volunteers. The second limitation is the observational nature of the study, which has meant that we have not had the opportunity to obtain a blood sample for all patients at all time points, due to the rapidity of the national vaccination programme in the UK. Nevertheless, as far as we are aware, this is the largest cohort of patients with mature B-cell Hodgkin and non-Hodgkin lymphoma for which antibody, cellular and pseudoneutralization data is available. As vaccine immune responses are heterogeneous, a large sample size is necessary to control for potentially confounding factors.

Whilst only a small cohort of patients with responses after the third vaccine dose have been included, similar trends are observed in vaccine response, and consistent with that reported by Greenberger et al(Greenberger et al., 2021a). The timing between vaccination and treatment completion continues to be a critical factor even with a third vaccine dose. Sixty-four per cent of participants vaccinated within 52 weeks of anti-CD20 exposure did not mount a detectable antibody response, suggesting that further doses within this time period is unlikely to be effective in inducing an antibody response and anti-SARS-CoV-2 monoclonal antibodies should be considered in these individuals. Similarly, minimal elevation was also observed in the anti-S IgG participants with HL who were vaccinated on chemotherapy. In contrast, it was reassuring that participants with indolent B-NHL demonstrated a rise in antibody levels after the third vaccine dose, indicating that those with ‘low’ antibody levels may benefit from a further dose of vaccine as part of their primary vaccination course.

In summary, we have demonstrated that the strongest predictor of antibody vaccine response in a large cohort of participants with lymphoid malignancies is the timing of treatment in relation to vaccination, regardless of the number of doses administered. These patients’ antibodies have the same functional ability as healthy participants to block viral spike protein binding to host ACE2 receptor, and vaccine cellular responses are preserved in most.

The exceptions are people with indolent B-NHL who may have impaired antibody levels regardless of treatment and also impaired cellular responses. Immunological monitoring of this cohort of individuals should be urgently implemented to guide the need for further vaccine doses, or alternative strategies such as prophylactic antibodies. Finally, those who receive B-cell depleting treatments such as anti-CD20, should be re-vaccinated with two doses of vaccine post-treatment.

## Data Availability

All data produced in the present study are available upon reasonable request to the corresponding author.

## ACKNOWLEDGEMENTS

The PROSECO study is funded by the Blood Cancer UK Vaccine Research Collaborative, which is led by Blood Cancer UK in partnership with Myeloma UK, Anthony Nolan and the British Society for Haematology (21009), and supported by Cancer Research UK Advanced Clinician Scientist Fellowship to S.H.L. (A27179), the Cancer Research UK/NIHR Southampton Experimental Cancer Medicine Centre (A25141), the NIHR Southampton Clinical Research Facility (Southampton Research Biorepository) and NIHR Southampton Biomedical Research Centre. D.G. receives support from the NIHR Great Ormond Street Biomedical Research Centre. We thank members of the Blood Cancer UK Vaccine Task Force for their advice. We sincerely thank all the participating sites’ research staff for their hard work and the research participants.

## AUTHOR CONTRIBUTIONS

S.H.L. conceived, supervised, investigated, administered, curated the data, wrote the original draft, and acquired funding; D.G. supervised, analysed and review and edited the manuscript; N.C. and B.S. curated, analysed and validated data; M.J. investigated and analysed data; D.J.P. administered, analysed and reviewed and edited the manuscript, A.K., D.J., A.H.T., C.G., T.W., A.R.C. and B.W. investigated and analysed data; K.R. administered, K.L. and J.B. designed the database; A.T.B., D.A., R.L., J.F., O.B., V.W., A.B.A-N., L.R., A.O., G.P.C., T.M., C.P.F., M.A. and P.W.M.J. investigated and reviewed and edited the manuscript., S.N.F. contributed to methodology and reviewed and edited the manuscript, A.J.D. investigated and acquired funding, and reviewed and edited the manuscript. All authors reviewed the manuscript.

## METHODS

### Experimental model and subject details

#### Patient data collection

This study was approved by the UK National Health Service Health Research Authority (IRAS 294739; 233768). It is a multicentre, prospective observational cohort study which enrolled participants from local hospital databases or outpatient clinics from January to September 2021, after informed consent. Eligible participants had to be 18 years old or older and have a confirmed diagnosis of a mature lymphoid malignancy. Information regarding demographic details, height and weight, disease characteristics, cancer treatment history, co- morbidities, COVID-19 infection status, medication history and SARS-CoV-2 vaccination details and adverse events were collected. Subjects also underwent peripheral blood sampling prior to vaccination, 4 weeks post first dose, 2 to 4 weeks post second dose, either 24 weeks post second dose or 6 weeks prior to the third dose and 4 to 8 weeks after the third dose.

Participants were recruited into four main diagnostic categories, HL, aggressive B-NHL, indolent B-NHL and PTCL based on their histological diagnosis, where available. Participants with nodular lymphocyte predominant Hodgkin lymphoma (NLPHL) were classified as indolent B-NHL for the purpose of this study due to the treatment of this disease with anti-CD20 monoclonal antibodies. participants were allocated into ‘no treatment’ vs ‘on treatment’ groups. Participants who had not received systemic anti-lymphoma therapy for their disease, or whose treatment completed 24 weeks prior to the first vaccine dose was allocated to the ‘no treatment’ group. Participants in the ‘on treatment’ group either received their first vaccine dose within 24 weeks of completing systemic anti-lymphoma therapy or who started systemic anti-lymphoma therapy within 4 weeks of the first vaccine dose. For the uniformity, one month is defined as 4 weeks in this study.

Healthy controls were vaccinees who received vaccine as part of the government rollout and donated serum post vaccination for essay evaluation through verbal consent.

### Method Details

#### Anti-SARS-CoV-2 S, RBD and N IgG Assay

Antibody quantification was undertaken using frozen serum or plasma in the WHO International Reference Laboratory for Pneumococcal Serology at University College London, London, UK. A multiplex electrochemiluminescent MSD assay (MesoScale Discovery) was performed as previously described with SARS-CoV-2 antigens N, RBD and trimeric S antigen (wild type Wuhan strain)(Johnson et al., 2020). The assay was calibrated with the WHO International reference serum (NIBSC 20/136) and results expressed as binding antibody units per ml (BAU/mL). Anti-N IgG concentration of 0.64 BAU/mL or lower, anti-RBD IgG of 0.73 BAU/mL or lower and anti-S IgG level of 0.55 BAU/mL or lower were below the detection limit. Subjects with an anti-N IgG level exceeding 6.60 BAU/mL were considered to have encountered previous SARS-CoV-2 and were excluded from primary analysis.

#### Pseudoneutralization (ACE2 receptor blocking) assay

Samples with detectable anti-S and anti-RBD IgG antibodies were evaluated by a pseudoneutralization assay as previously described in the WHO International Reference Laboratory for Pneumococcal Serology at University College London, London, UK (Johnson et al., 2020). Briefly, a standard curve derived from doubling dilutions of an anti-S monoclonal antibody as well as control or test sera (1:10 dilution) were added to a 96-well MULTI-SPOT® plates coated with S or RBD antigen. After incubation, ACE2 conjugated MSD SULFO-TAG was added and the plates read using a MESO® SECTOR S600 reader. The amount of ACE2 receptor blocking was calculated from the standard curve for each unknown serum and expressed as a titer. Samples that were above the maximum for the standard curve were assigned a titer of 2.5 times the maximum (500).

#### SARS-CoV-2 IFNγ Enzyme linked Immunospot (ELISpot) assay

SARS-CoV-2-specific T-cell responses were assessed using frozen PBMCs in a standardised ELISpot assay at the WISH laboratory, Southampton, UK (a Good Clinical Practice (GCP) compliant laboratory). Briefly, ELISpot plates were coated with anti-human IFNγ antibody overnight. The following day, thawed cells were resuspended at 4 × 10^6^ cells/ml in Roswell Memorial Park Institute (RMPI) media supplemented with 10% human AB serum, sodium pyruvate and L-glutamine and rested for 2 to 4 h at 37°C, 5% CO_2_. The cells (approx. 4×10^5^ per well [1×10^5^ for PHA wells]) were plated in triplicate wells and incubated alone or with 1 µg/ml SARS-CoV-2 PepMix™ peptide pools (JPT) (peptide spanning the spike glycoprotein provided as 2 separate peptide pools) or 5 µg/ml phytohaemagglutinin (Sigma) for 18-20 h. After incubation, PBMCs and peptides were washed off and plates were incubated with biotinylated anti-human IFNγ (7-B6-1, Mabtech) for 90 min at 37°C, 5% CO_2_. The plates were then washed with PBS-Tween four times before incubation with streptavidin alkaline phosphatase (Mabtech) for 60 min at 37°C, 5% CO_2_. After 4 further washes with PBS-Tween, alkaline phosphatase chromogenic substrate (Novex) was added, spots allowed to develop for ≤10 minutes, and plates read on an ELISpot plate reader (Autoimmun Diagnostika GmBH). Spots were assessed using ELISpot 6.0 software and results reported as spot forming units per 10^6^ cells (SFU/10^6^ cells). The mean of triplicate unstimulated wells was subtracted from individual replicate stimulated wells (for both peptide pool 1 and 2). The corrected values for each peptide pool were then summed, and the mean of the total peptide pool response was calculated. The test result is considered positive if it is more than 28.06 SFU/10^6^ cells. This value represents the upper 95% confidence interval limit of 32 pre-vaccination samples.

#### Peripheral blood T, B and NK cell quantification

Enumeration of peripheral blood T, B and NK cells were undertaken using multicolour flow cytometry as previously described(Turaj et al., 2017). Briefly, thawed PBMCs were suspended in staining buffer and fluorescent conjugated antibodies added for 15 min, room temperature prior to two washes, 4% paraformaldehyde fixation and collection on the flow a FACSCanto (BD Biosciences). Data was analysed using Cytobank (Cytobank).

PBMCs were analysed for B cells (anti-CD19-PE-Cy7, anti-CD3-FITC), CD4^+^ T cells (anti-CD3-FITC, anti- CD4-BV510), CD8^+^ T cells (anti-CD3-FITC, anti-CD8-APC eF780) and NK cells (anti-CD3-FITC, anti-CD56- PE) and markers of activation (HLA-DR-PerCP-Cy5.5, CD38 APC). Counting beads (Biolegend) were used as per manufacturer’s instructions for cell counting.

### Statistical analysis

Patient characteristics were summarised using descriptive statistics appropriate to the distribution. The association between more than two continuous variables were tested using Kruskal-Wallis Rank Sum test. Where only two groups exist, a Mann-Whitney test was used. For binary outcomes, a logistic regression model was used for both univariable and multivariable analyses and odds ratios reported with 95% confidence intervals. All p values less than 0.05 was considered statistically significant. All analyses were done in Stata, version 16.0 and GraphPad Prism, version 9.

