## Supplemental Data for "Humoral and cellular responses to SARS-CoV-2 vaccination in patients with lymphoid malignancies"

### Supplemental Tables

| | OR favoring undetectable anti-S IgG (95% CI) |
|---|---|
| **Treatment group** | |
| - **On** | **10.96 (6.44, 18.63; p<0.001)** |
| - **No** | Reference |
| **Vaccine** | |
| - **ChAdOx1** | Reference |
| - **BNT162b2** | 0.96 (0.60, 1.52; p=0.854) |
| **Disease group** | |
| - **Aggressive BNHL** | Reference |
| - **HL** | **0.17 (0.05, 0.58; p=0.005)** |
| - **Indolent BNHL** | 1.52 (0.93, 2.50; p=0.098) |
| - **Peripheral TNK** | 1.59 (0.46, 5.52; p=0.464) |
| **Gender** | |
| - **Male** | Reference |
| - **Female** | 0.93 (0.59, 1.47; p=0.758) |
| **Anti-CD20** | |
| - **No** | Reference |
| - **Yes** | **13.72 (7.78, 24.22; p<0.001)** |
| **Age** | **1.04 (1.02, 1.05; p<0.001)** |
| **Disease remission status** | |
| - **Complete/partial** | Reference |
| - **Stable disease** | 0.77 (0.36, 1.67; p=0.513) |
| - **Progressive disease** | 2.10 (0.76, 5.86; p=0.155) |
| **Serum IgG status** | |
| - **Normal/High** | Reference |
| - **Low** | **3.32 (1.62, 6.77 ; p=0.001)** |

**Table S1. Univariable analysis of factors contributing to undetectable anti-S IgG**
Univariable logistic regression reporting odds ratios and 95% confidence intervals. Older age, anti-CD20 treatment in the last 12 months, low serum IgG status and being in the 'on treatment' group were associated with higher risk of undetectable anti-S IgG.

| | **OR favoring undetectable anti-S IgG (95% CI)** |
|---|---|
| **Treatment group** | |
| - **On** | **7.22 (3.80, 13.73; p<0.001)** |
| - **No** | REF |
| **Vaccine** | |
| - **ChAdOx1** | REF |
| - **BNT162b2** | 0.89 (0.49, 1.62; p=0.695) |
| **Disease group** | |
| - **Aggressive B-NHL** | REF |
| - **HL** | 0.70 (0.16, 3.08; p=0.641) |
| - **Indolent B-NHL** | 1.53 (0.80, 2.92; p=0.202) |
| - **PTCL** | 2.60 (0.53, 12.76; p=0.240) |
| **Gender** | |
| - **Male** | REF |
| - **Female** | 0.82 (0.45, 1.50; p=0.525) |
| **Anti-CD20 administered in last 12 months** | |
| - **No** | REF |
| - **Yes** | **5.60 (2.73, 11.52; p<0.001)** |
| **Age** | **1.03 (1.01, 1.06; p=0.011)** |
| **Disease remission status** | |
| - **Complete/partial** | REF |
| - **Stable disease** | 0.60 (0.26, 1.85; p=0.468) |
| - **Progressive disease** | 2.44 (0.62, 9.51; p=0.200) |

**Table S2. Multivariable analysis of factors contributing to undetectable anti-S IgG**
Multivariable logistic regression model, reporting odds ratios and 95% confidence intervals. Older age, anti-CD20 treatment in the last 12 months and being in the 'on treatment' group were associated with higher risk of undetectable anti-S IgG.

| | **OR favoring a positive IFNγ ELISpot response** |
|---|---|
| | OR (95% CI; p-value) |
| **Treatment group** | |
| - **On** | 2.17 (0.99, 4.72; p=0.051) |
| - **No** | REF |
| **Vaccine** | |
| - **ChAdOx1** | **2.01 (1.06, 3.79; p=0.031)** |
| - **BNT162b2** | REF |
| **Disease group** | |
| - **Aggressive BNHL** | REF |
| - **HL** | 0.86 (0.26, 2.78; p=0.797) |
| - **Indolent BNHL** | 0.64 (0.31, 1.34; p=0.238) |
| **Gender** | |
| - **Male** | REF |
| - **Female** | 0.67 (0.35, 1.28; p=0.230) |
| **Anti-CD20** | |
| - **No** | REF |
| - **Yes** | 0.47 (0.20, 1.08; p=0.076) |
| **Age** | 1.00 (0.97, 1.02; p=0.774) |
| **Disease remission status** | |
| - **Complete/partial** | REF |
| - **Stable disease** | 1.13 (0.37, 3.40; p=0.834) |
| - **Progressive disease** | 0.75 (0.18, 3.16; p=0.698) |

**Table S3. Multivariable analysis of factors favoring reduced cellular responses**
Multivariable logistic regression model, reporting odds ratios and 95% confidence intervals. ChAdOx1 vaccination vaccine was the only significant predictor of positive IFNγ ELISpot response.

**Supplemental Figure Legends**

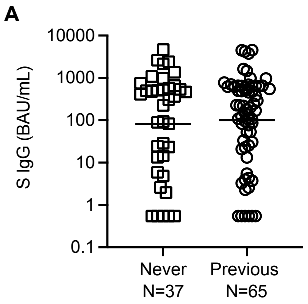


**Figure S2, related to Figure 2**

Anti-S IgG levels in participants with indolent B-NHL are similar between those who had received previous anti-systemic therapy compared to those who had never received systemic therapy. GMC shown, Kruskal-Wallis test.

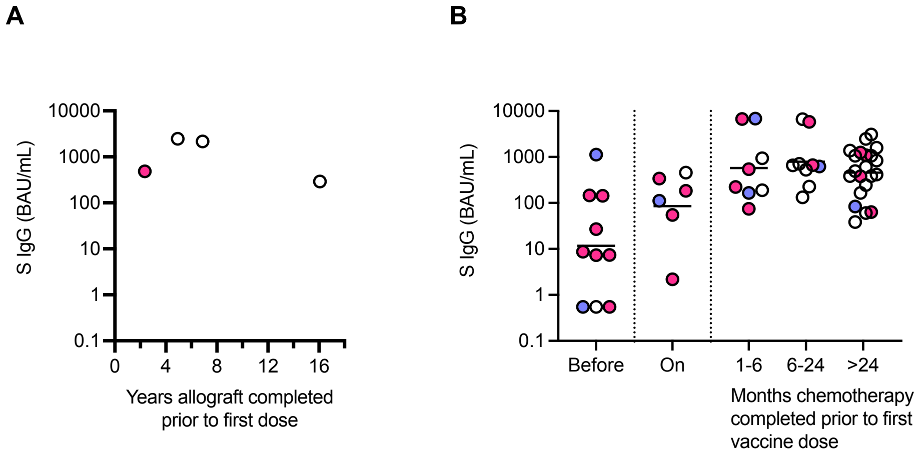


**Figure S3, related to Figure 3**

(A) Antibody responses are preserved allogeneic stem cell allograft recipients transplanted >2 years previously.
(B) Antigen-specific T-cell responses are detectable in participants with HL regardless of timing of chemotherapy. Positive test (pink), blue (negative) and white (not tested).

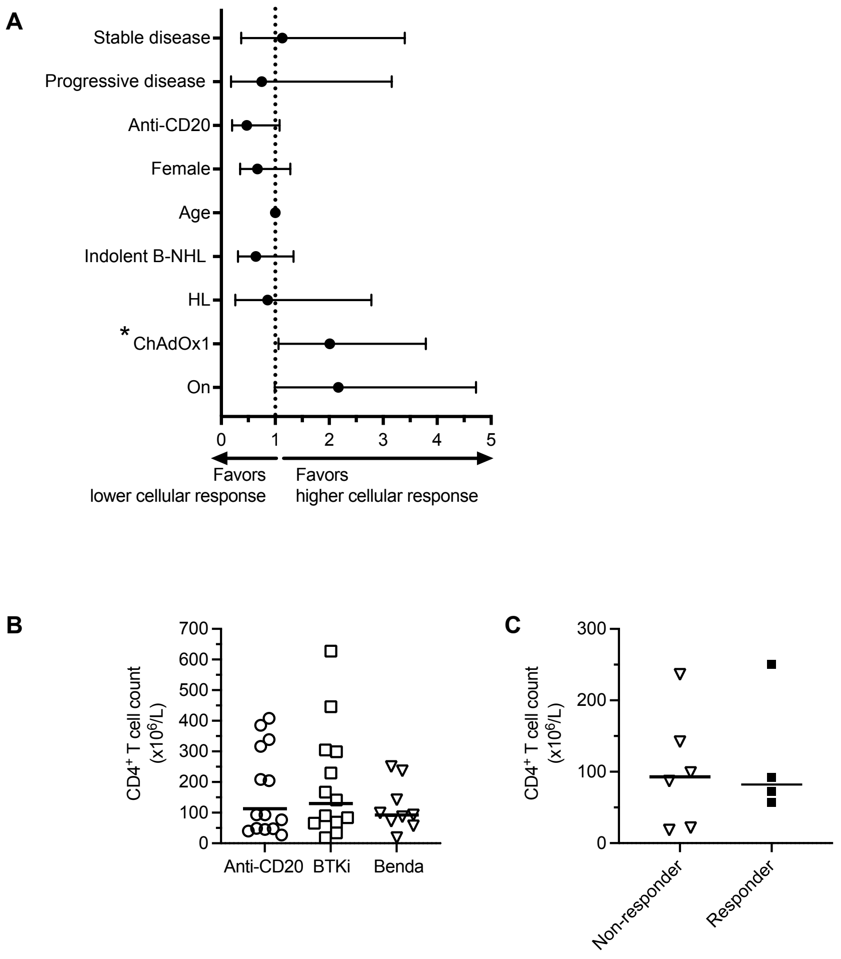


**Figure S4, related to Figure 4**

(A) ChAdOx1 vaccination is associated with a lower cellular response on multivariable logistic regression analysis. *implies statistical significance.

(B) No difference in CD4+ T cell counts between participants on or recently treated with anti-CD20, BTKi or bendamustine.

(C) No difference in CD4+ T cells was observed in bendamustine-exposed cases who did not have an antigen-specific T-cell response (non-responder) compared to those who had a positive test (responder)
